# Assessing the clinical significance of a novel rare variant in Loeys-Dietz Syndrome by combining AI-driven modelling and cell biology

**DOI:** 10.64898/2026.03.30.26349510

**Authors:** Nihad Boukrout, Clément Delage, Thomas Comptdaer, Wendy Arondal, Amine Jemel, Nadia Azabou, Mouna Bousnina, Molka Mallouki, Naoual Sabaouni, Rihab Arbi, Salma Kchaou, Haithem Ammar, Saoussen Hantous-Zannad, Houweyda Jilani, Yasmina Elaribi, Lamia Benjemaa, Cynthia Van der Hauwaert, Romain Larrue, Meyling Cheok, Michaël Perrais, Bruno Lefebvre, Christelle Cauffiez, Nicolas Pottier

**Affiliations:** Univ. Lille, CNRS, Inserm, CHU Lille, Institut Pasteur de Lille, UMR9020-U1277-CANTHER-Cancer Heterogeneity Plasticity and Resistance to Therapies, Lille, Hauts-de-France 59000, France; Service de Toxicologie et Génopathies, CHU Lille, Lille, Hauts-de-France 59000, France; Cardiovascular surgery department, Abderrahmen Mami hospital, Ariana 2081, Tunisia; Faculty of Medicine of Tunis, University of Tunis El Manar, Tunis 1007, Tunisia; Pediatric cardiac surgery department, Bechir Hamza Children’s hospital, Tunis 1006, Tunisia; Medical imaging department, Abderrahmen Mami hospital, Ariana, 2081, Tunisia; Research laboratory LR20SP02 “cardio-thoracic imaging”, Abderrahmen Mami hospital, Ariana, 2081, Tunisia; Genetic department, Mongi Slim hospital, La Marsa (Sidi Daoud), Tunis 2046, Tunisia; Research laboratory LR22SP01 “mother-child health”, Mongi Slim hospital, La Marsa (Sidi Daoud), Tunis 2046, Tunisia; Research laboratory LR16SP01 “infertility and oncofertility”, Aziza Othmana hospital, Tunis 1008, Tunisia

**Keywords:** Loeys–Dietz syndrome, genetic variant, structural modelling, TGF-β signalling

## Abstract

Loeys–Dietz syndrome (LDS) is an autosomal dominant connective-tissue disorder caused by genetic variants in TGF-β pathway genes, most often *TGFBR1/2*. While pathogenic *TGFBR2* mutations usually cluster in the kinase domain and disrupt SMAD signaling, the real challenge in accurate genetic testing is separating the variants that truly affect *TGFBR2* function from the rare benign alterations that only look suspicious at first glance. Therefore, there is a pressing need to develop methods that can improve functional variant interpretation. Here, we describe and characterize the functional impact of a novel genetic variant in the TGFBR2 kinase domain (E431K), in a patient with the clinical diagnosis of syndromic genetic aortopathy. We assessed the structural and functional consequences of this variant using AI-driven molecular modelling and *in vitro* cell-based assays. A high-quality homology-based model of TGFBR2 was generated and computational mutagenesis based on the structural context and evolutionary conservation was used to forecast variant pathogenicity. Relative to wild type, the variant affects protein stability by disrupting intramolecular interactions and likely induces conformational changes that may affect kinase activity and thus TGF-β signalling. This was experimentally confirmed by showing abnormal protein level and alteration of canonical TGF-β pathway activation. Overall, our results establish that the E431K variant leads to aberrant TGF-β signalling and confirm the diagnosis of Loeys–Dietz syndrome type 2 in this patient.

## Introduction

Rare diseases, of which approximately 80% are genetic in origin, affect an estimated 3.5-5.9% of the global population and corresponding to nearly 300-400 million individuals worldwide. Although each disease is individually uncommon, their collective impact represents a significant global health burden (The Lancet Global Health 2024; Global Genes, 2023). With the advent of next-generation sequencing technologies, the ability to detect genetic variants has dramatically improved, offering not only higher diagnostic yield but also insights into the familial inheritance of disease and opportunities for personalized care (Schobers et al. 2024; Popova and Carabetta 2025). To date, hundreds of millions of genetic variants have been catalogued in public databases such as ClinVar and gnomAD (Landrum et al. 2020; Karczewski et al. 2020). However, most of them remain classified as variants of uncertain significance, underscoring the urgent need for functional studies and careful clinical correlation to establish their pathogenicity (Larrue et al. 2020, 2024; Park et al. 2025; Ma et al. 2025). This represents one of the main challenges currently facing medical genetics: moving from simply detecting variants to definitively interpreting and translating them into clinical practice.

Among rare genetic disorders, the Loeys–Dietz syndrome (LDS) (OMIM:610168) is a particularly severe connective tissue disorder first described in 2005 (Loeys et al. 2005). LDS manifests with arterial aneurysms and dissections, frequently accompanied by widespread arterial tortuosity, in association with characteristic craniofacial abnormalities (hypertelorism, bifid uvula, or cleft palate) and skeletal manifestations (pectus deformities, scoliosis, joint laxity) (Loeys et al. 2005, 2006). LDS, while genetically heterogeneous, is most commonly caused by pathogenic variants in genes encoding components of the transforming growth factor-beta (TGF-β) signalling pathway, including *TGFBR1*, *TGFBR2*, *SMAD2*, *SMAD3*, *TGFB2*, and *TGFB3* (Loeys et al. 2005, 2006; MacCarrick et al. 2014). TGFBR2 (TGF-β receptor type II), an enzyme-linked receptor with dual specificity of serine/threonine kinase and tyrosine kinase, plays a pivotal role in initiating TGF-β signalling (Lin et al. 1992). Indeed, upon ligand binding, TGFBR2 recruits and phosphorylates TGFBR1, which in turn activates downstream SMAD proteins to mediate canonical transcriptional response (Massagué 2012) (Fig. 1). Dysregulation of this pathway is not only central to the vascular and the connective tissue pathology observed in LDS, but also in the pathogenesis of other heritable connective tissue disorders, including Marfan syndrome or Shprintzen–Goldberg syndrome (Neptune et al. 2003; van Steensel et al. 2008; Stheneur et al. 2008). These TGF-β-related connective tissue diseases follow an autosomal dominant mode of inheritance, share significant phenotypic overlap, and have multisystem involvement with in particular cardiovascular complications such as arterial dissections, aneurysms, and valve prolapse (De Backer et al. 2013; Verstraeten et al. 2016; Takeda et al. 2018).

**Figure 1.**
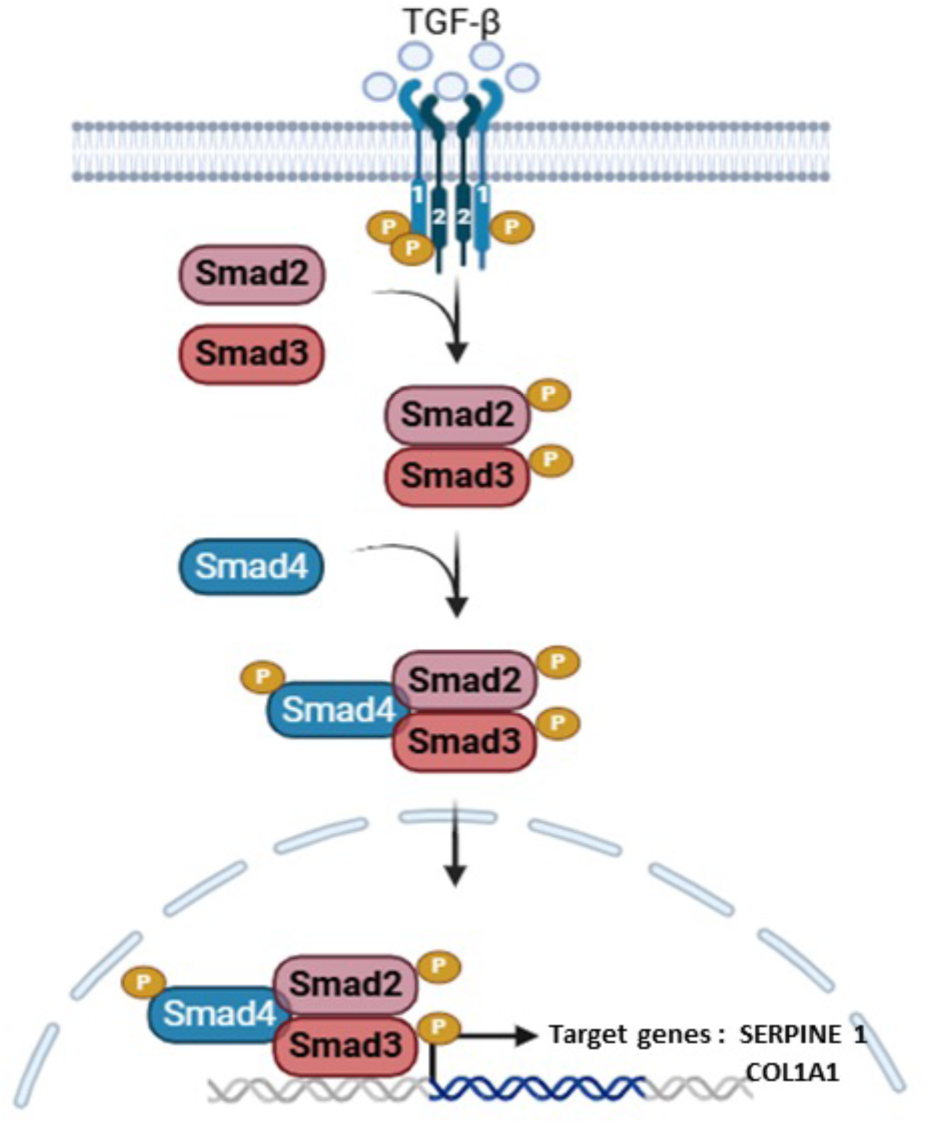
Canonical TGF-β signalling pathway. TGF-β ligand binds to the type II receptor (TGFBR2), which recruits and phosphorylates the type I receptor (TGFBR1). Activated TGFBR1 phosphorylates receptor-regulated SMADs (SMAD2/3), which form a complex with the common mediator SMAD4. This complex translocates to the nucleus, where it regulates the transcription of target genes controlling proliferation, differentiation, apoptosis, and extracellular matrix homeostasis

Although numerous variants of the *TGFBR2* gene are stored in genetic banks, only a limited number of them have undergone functional evaluation to clarify their causal role in pathogenesis of LDS (Cousin et al. 2017). Notable examples include the p.Val419Leu variant which impairs canonical TGF-β/SMAD signalling in cell-based assays (Cousin et al. 2017). In addition, previous systematic studies have revealed that multiple LDS-associated *TGFBR2* mutations exert a dominant-negative effect and diminish SMAD/ERK activation (Horbelt et al. 2010). Altogether, these studies highlight the need to combine genomic sequencing with orthogonal functional testing and clinical correlation in order to discriminate genuine pathogenic variants from rare neutral mutation.

## Results

### Case Presentation: clinical findings and results of the genetic testing

The novel missense E431K variant in the *TGFBR2* gene identified in this study was detected by genetic testing of a male patient (age of 45-50 years) referred to the Mongi Slim Hospital (Tunisia) for suspected hereditary connective tissue disorders (Fig. 2). To investigate the genetic basis of the patient’s condition, whole-exome sequencing was undertaken. This analysis revealed 83 candidate variants (i.e. located within protein-coding regions) distributed across 81 genes known to be associated with genetic diseases (Table 1). Two rare missense variants affecting *TGFBR2* (NM_003242.6:c.1291G>A) and *FLNA* (NM_001110556.2:c.7450C>T, rs782557713) were further considered given the clinical diagnosis of syndromic genetic aortopathy (Table 1 and Fig. 3a). The hemizygous rs782557713 variant is known and broadly considered likely benign in multiple ClinVar entries. In contrast, the heterozygous *TGFBR2* variant has not been reported to date. It is absent from population reference datasets such as gnomAD, which usually suggests a non-disease role, but its predicted functional impact based on metapredictors such as REVEL and MetaLR remains inconclusive and MetaLR. All remaining variants were categorized as benign/likely benign, variants of uncertain significance, or single heterozygous loss of function alleles in recessive genes (e.g., *UBR1*, *NEB*, *CUL7*) unrelated to patient’s condition. There were consequently considered non-contributory. Therefore, the NM_003242.6(*TGFBR2*):c.1291G>A mutation was the most likely disease-causal variant and additional analyses and experiments were performed to determine its clinical significance.

**Figure 2.**
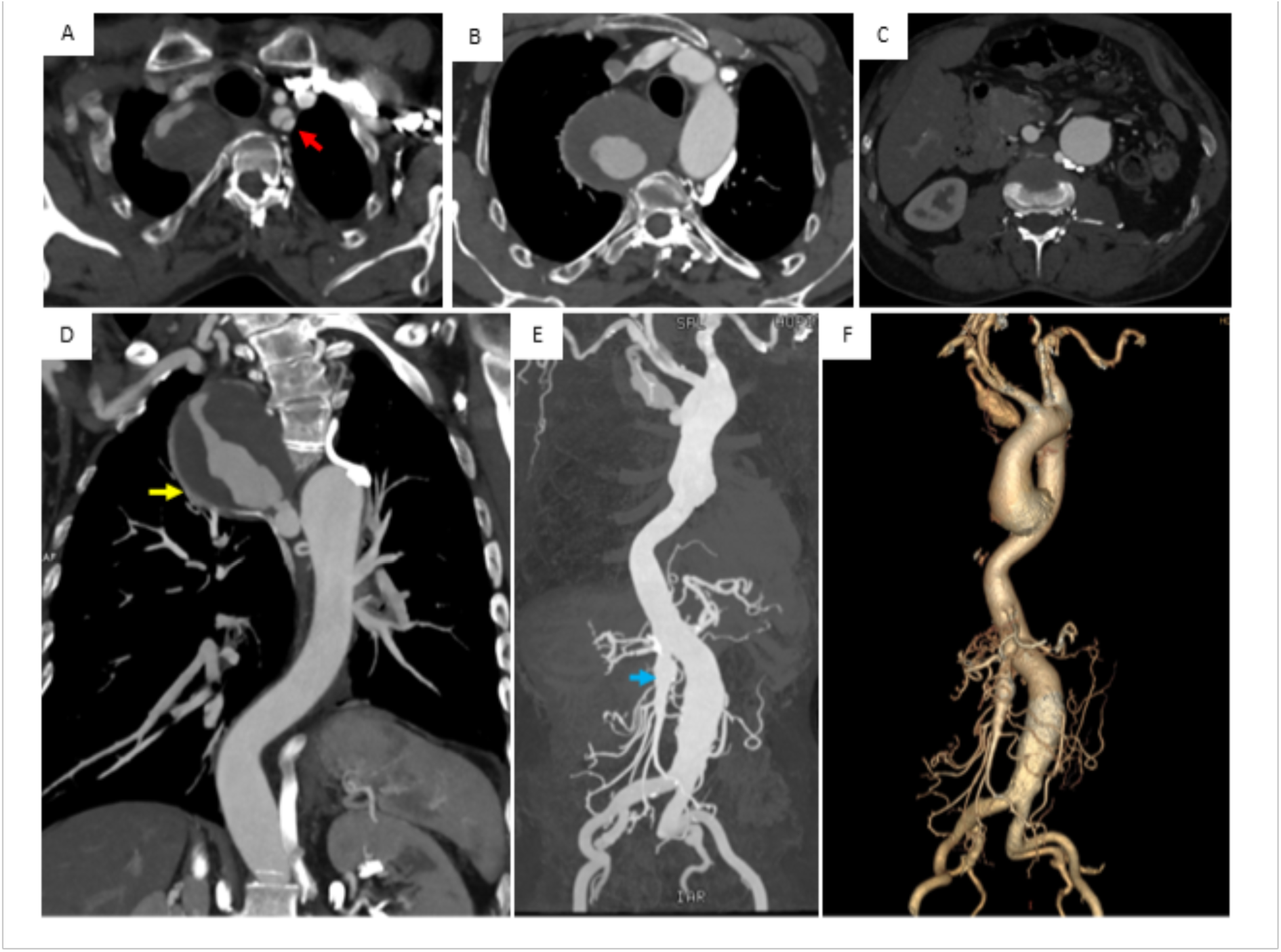
Proband vascular imaging. (a-f) Aortic computed tomography angiography of the patient: (a) Cross section showed a left subclavian dissection which is dilated (red arrow), (b) Large partially thrombosed aneurysm of the 6th right intercostal artery (c) Fusiform aneurysm of the supra renal abdominal aorta (d,e) Coronal 2D and maximum intensity projection reconstruction show multiple arterial aneurysms Yellow arrow : partially thrombosed aneurysm of the 6th intercostal artery Blue arrow: superior mesenteric artery aneurysm (f) Volume rendering (VR) three-dimensional reconstructed images show multiple arterial aneurysms affecting the left subclavian artery, the 6th right intercostal artery, the superior mesenteric artery and the abdominal aorta.

**Figure. 3.**
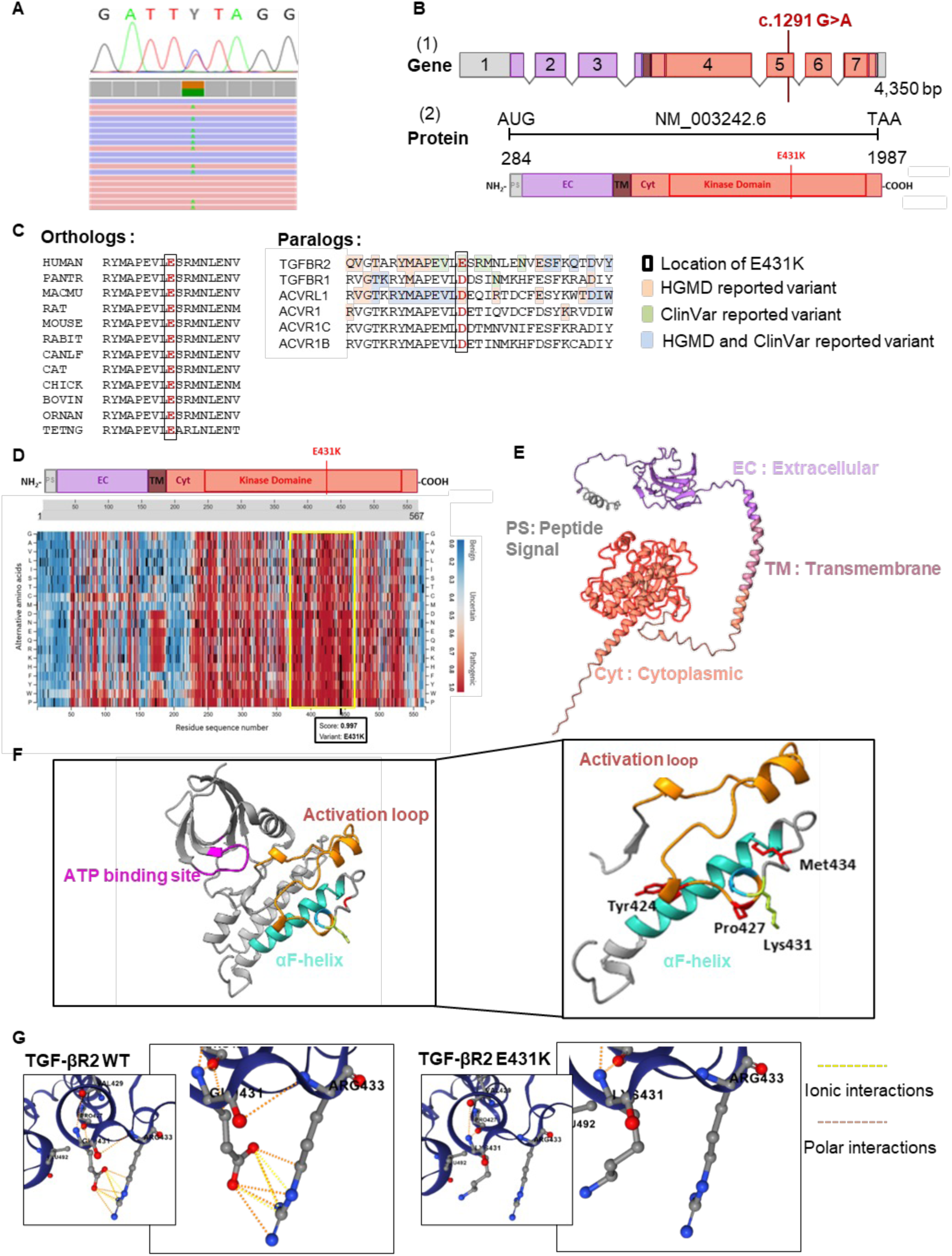
Identification and characterization of the TGFBR2 E431K variant. (a) IGV screenshot showing the heterozygous NM_003242.6(*TGFBR2*):c.1291G>A mutation identified by whole-exome sequencing and its subsequent confirmation by Sanger sequencing. (b-1) Schematic representation of *TGFBR2* gene structure. (b-2) Domain structures of TGFBR2 highlighting the position of the E431 residue within the serine/threonine kinase domain. (c) Multiple sequence alignment (MSA) of vertebrate orthologs and paralogous proteins generated with Clustal Omega; missense variants reported as pathogenic in HGMD and ClinVar are highlighted. (d) AlphaMissense pathogenicity scores for residues surrounding the E431 residue. (e) high-quality structural model of TGFBR2 generated by AlphaFold3. (f) Mapping of residue E431 within the kinase domain. (g) DDMut-predicted impact of the E431K substitution on TGFBR2 stability

**Table 1:**
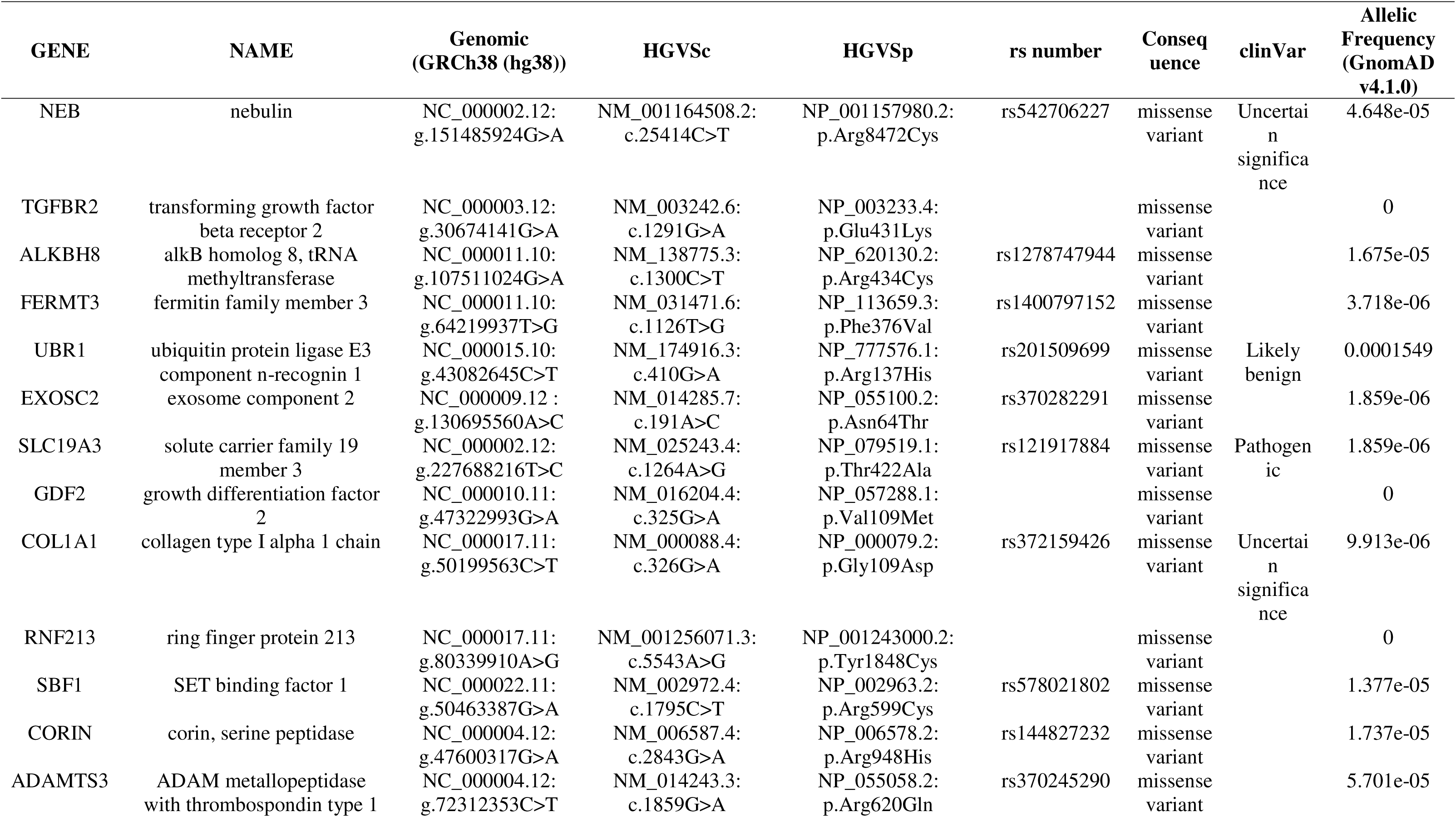
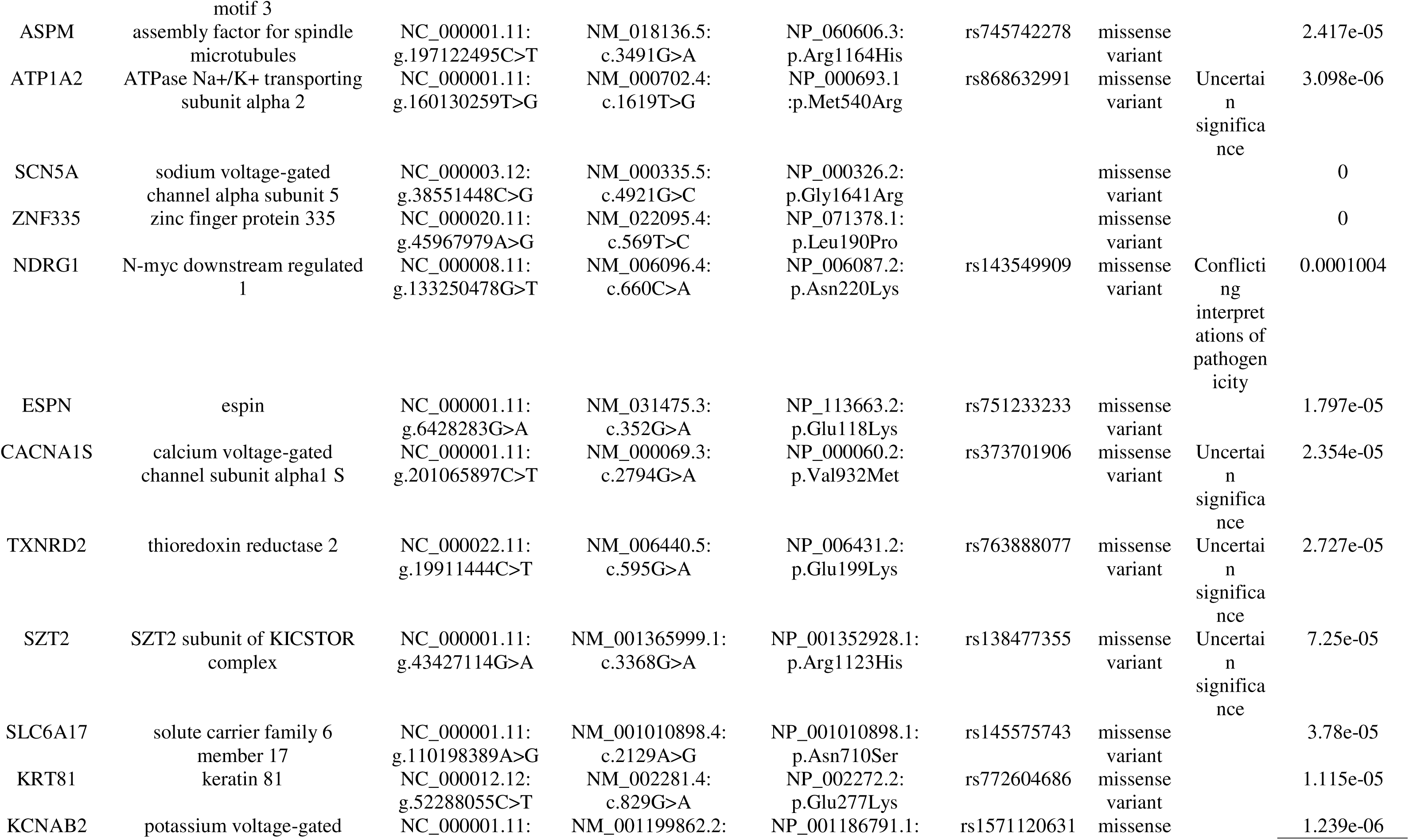
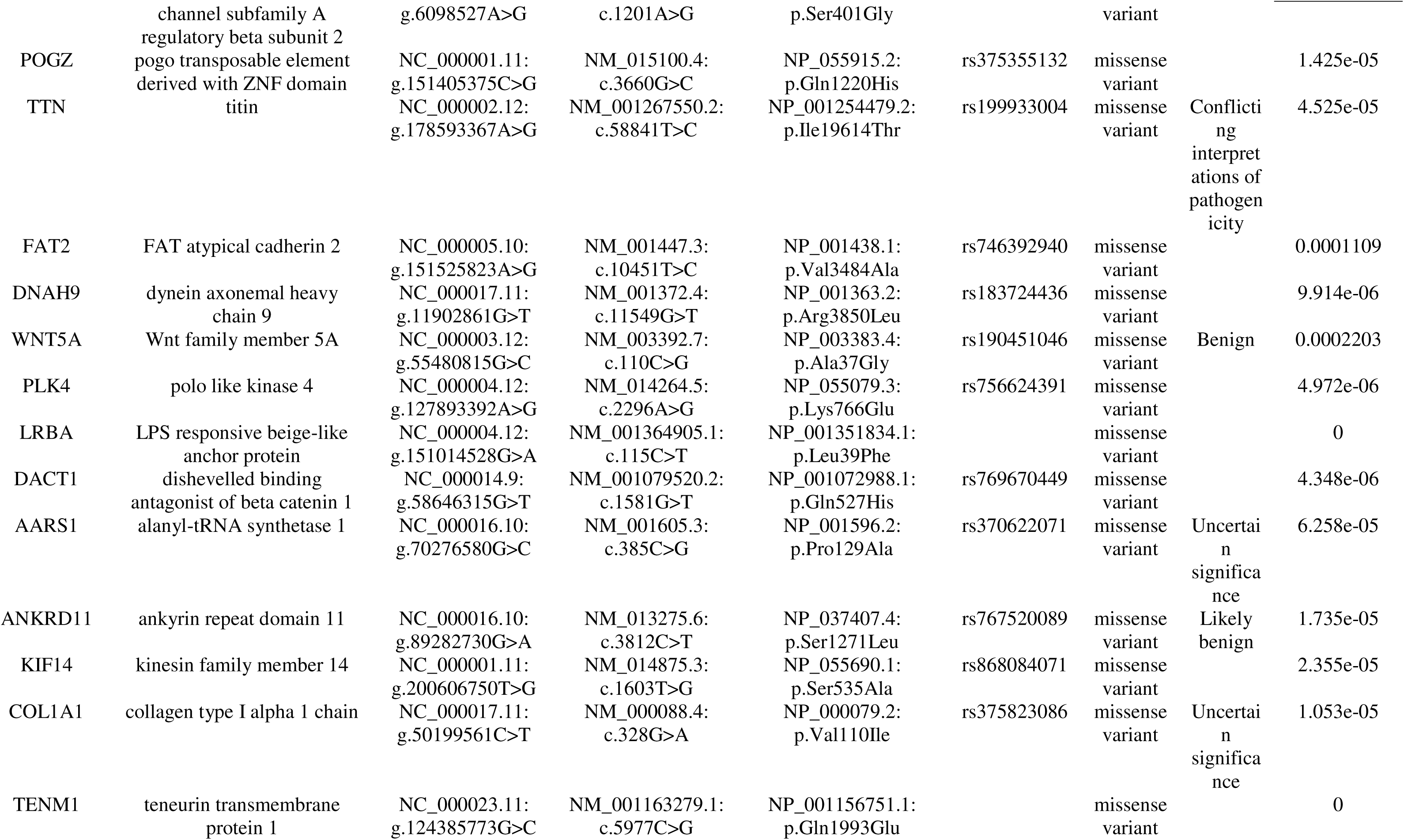
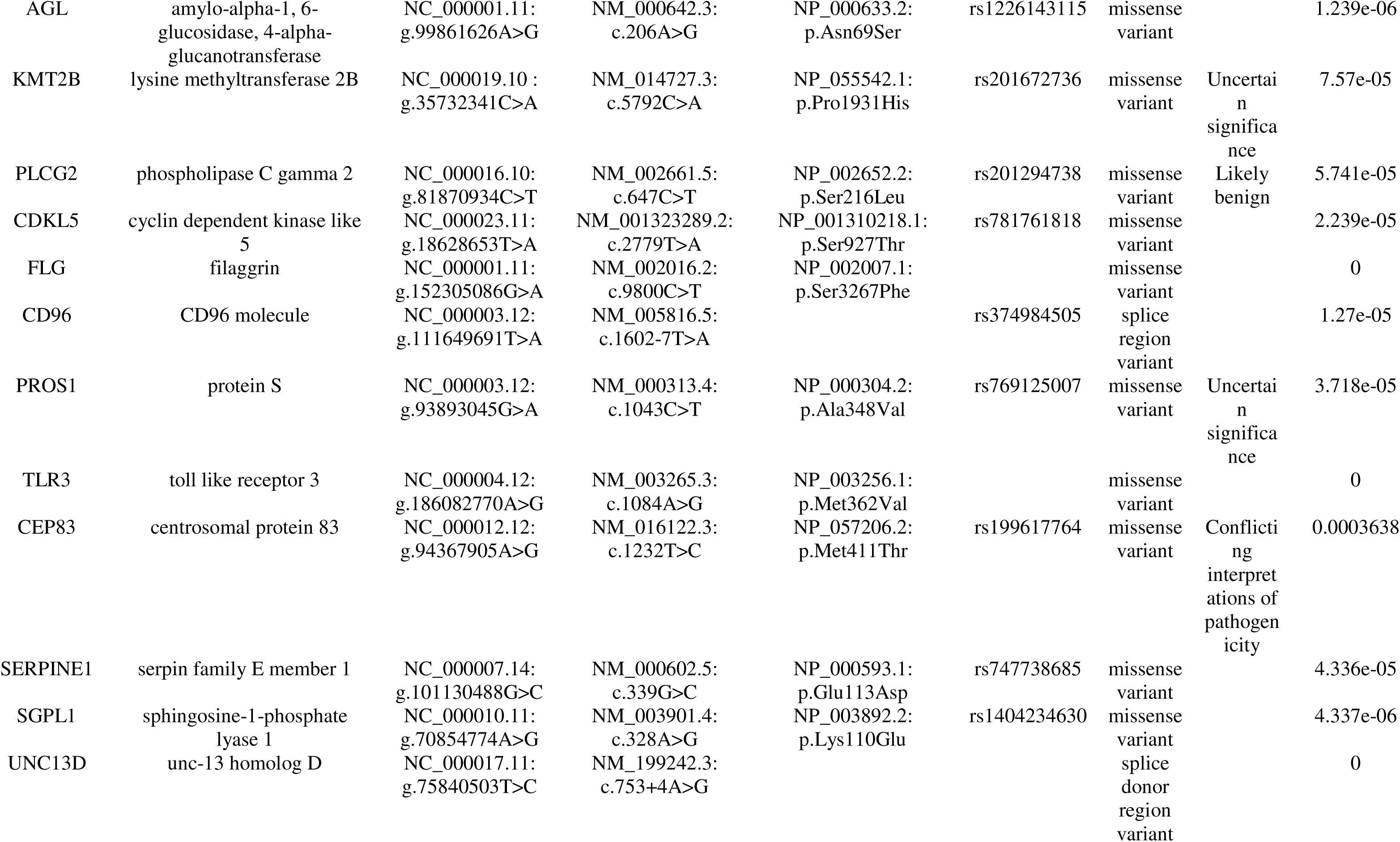
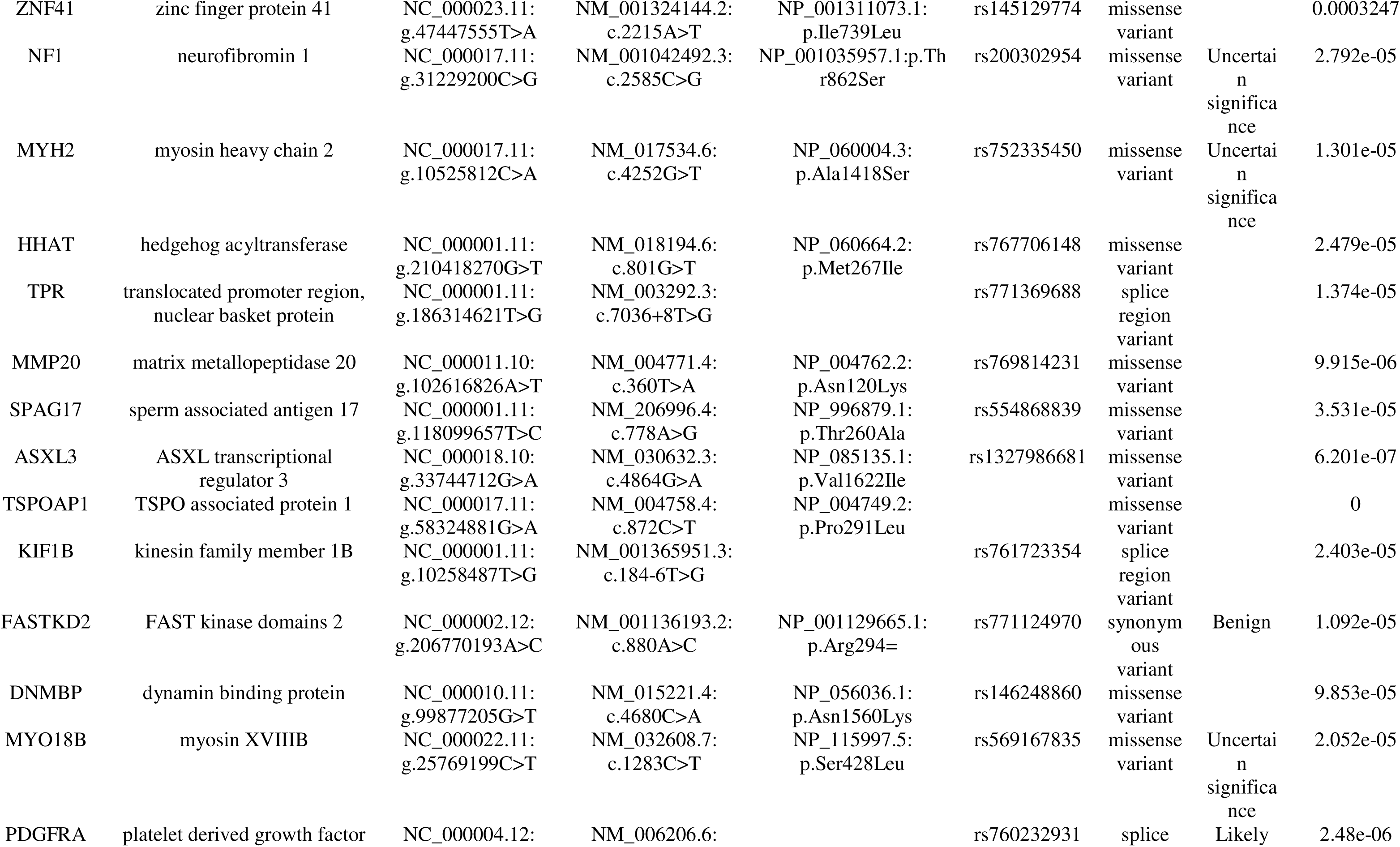
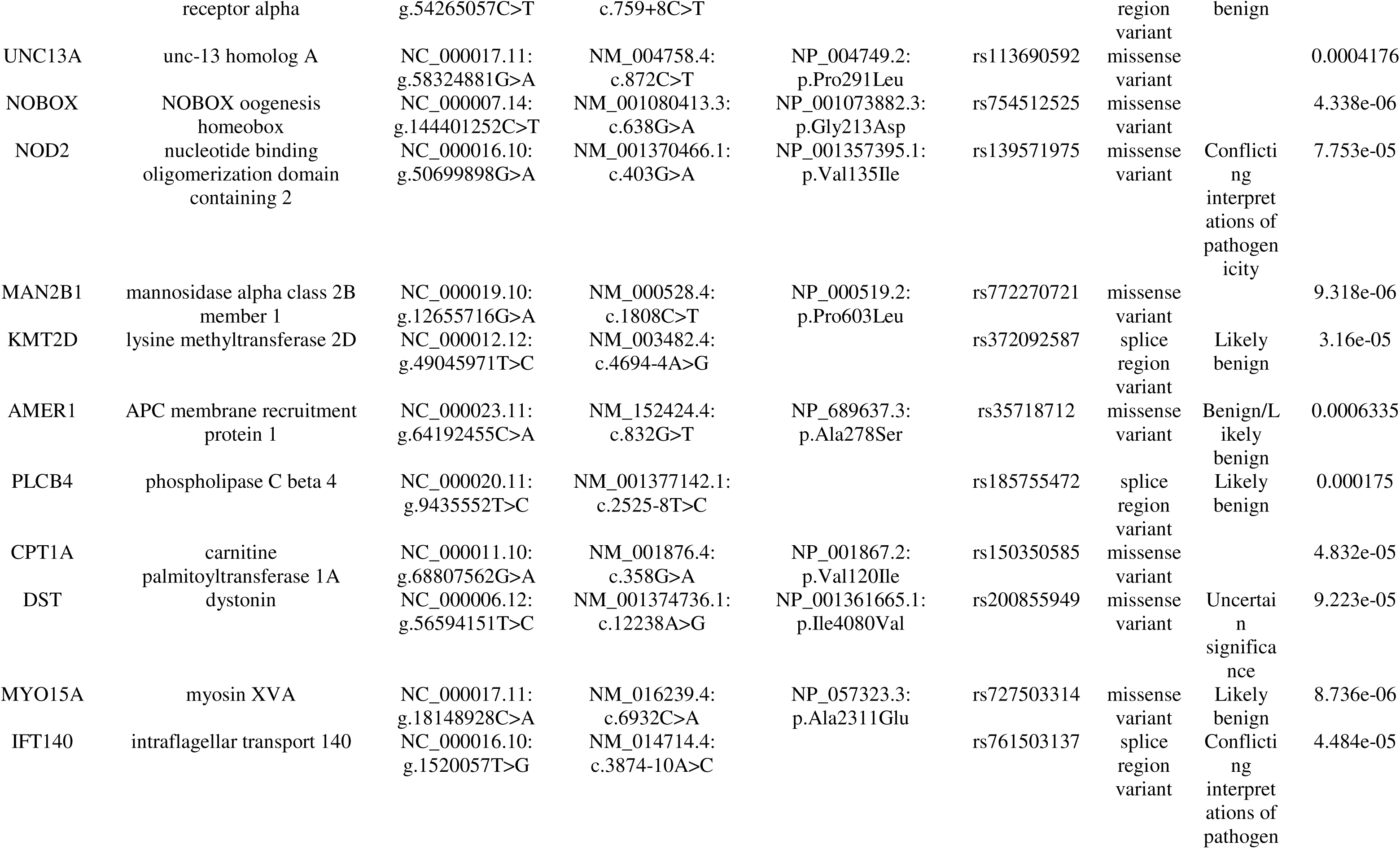
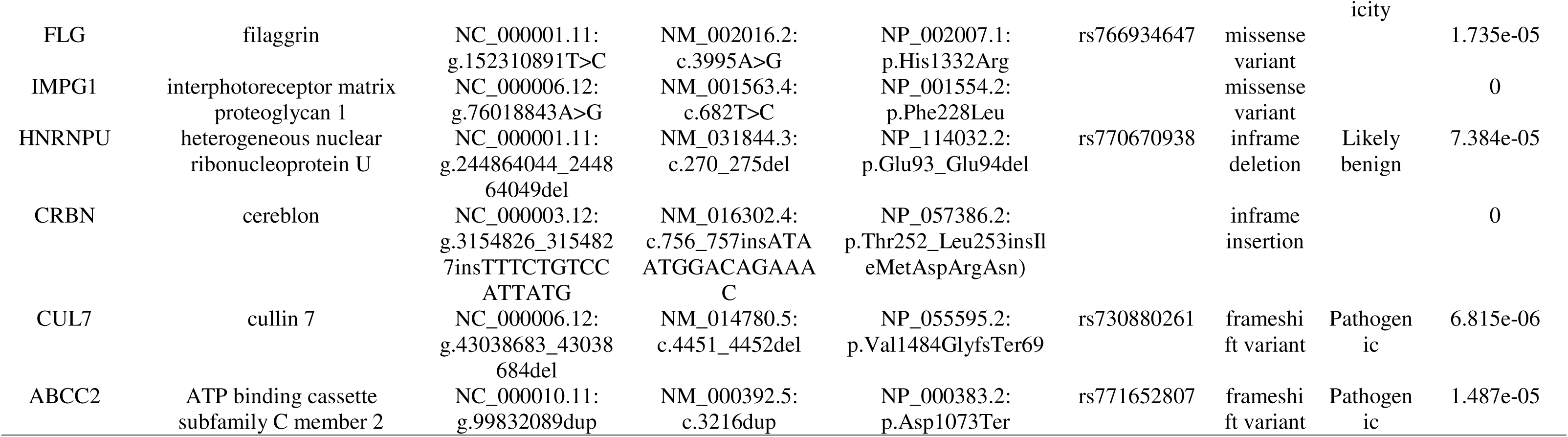
Candidate variants identified by whole-exome sequencing.

### AI-driven structural modelling of the E431K TGFBR2 variant

Sanger sequencing confirmed the heterozygous NM_003242.6(*TGFBR2*):c.1291G>A substitution in exon 5 of the gene (Fig. 3a, 3b-1), resulting in a missense change within the protein kinase domain at position 431 of the TGFBR2 protein (p.Glu431Lys and thereafter referred to as E431K) (Fig. 3b-2). Interestingly, this glutamic acid residue is strongly conserved across vertebrates and a negatively charged amino acid (glutamic acid or aspartic acid) is consistently found at this position in paralogs (Fig. 3c). As known disease-associated neighbouring missense changes are reported in the Human Gene Mutation Database (Stenson et al. 2014) or ClinVar (Landrum et al. 2020), we also mapped these pathogenic variants onto the paralog multiple sequence alignment. This analysis showed that known deleterious variants are located in the region surrounding the 431 position across several paralogs (Fig. 3c). Interestingly, nearly all single amino acid changes in this region have high AlphaMissense-predicted pathogenicity scores (Cheng et al. 2023), and the present variant was classified by this algorithm as likely pathogenic (score of 0.997) (Fig. 3d). To gain more insights into the functional impact of the E431K variant, we first generated a high-quality homology-based model of TGFBR2 using AlphaFold3 (Abramson et al. 2024), which unambiguously mapped the E431 residue within the serine/threonine kinase domain (Fig. 3e). We next aimed to refine the structural context of this residue and to map the neighbouring motifs most likely to be perturbed by the E431K mutation. AlphaFold3 modelling positioned the residue within a structurally constrained region adjacent to elements essential for preserving the architecture and catalytic competence of the TGFBR2 kinase domain. More precisely, this residue is spatially located nearby the αF helix, a conserved α-helix within the C-lobe of the kinase domain that forms part of the structural core of the kinase fold and helps maintaining the architecture of the catalytic site (Zhang et al. 2012), and the autophosphorylation site Y424 required for TGFBR2 kinase activity (Lawler et al. 1997). Nous The domain segment containing this variant also includes two ClinVar-reported pathogenic substitutions (P427L (Loeys et al. 2006) and M434K Richards et al. 2015), further underscoring the importance of this region (Fig. 3f). Given this structural context, the impact of E431K on protein stability was then assessed using the DDMut tool (Zhou et al. 2023), which predicted destabilization of the protein (ΔΔG stability (wt→mt) = −0.67 kcal/mol), due to the loss of polar and ionic interactions established with the arginine residue in position 433 of the TGFBR2 kinase domain (Fig. 3g). Then, we assessed robustness by reapplying DDMut to well characterized 419L variant; the model again indicated destabilization (ΔΔG (wt→mt) = −0.21 kcal/mol), consistent with our interpretation (data not shown).

#### Functional impact of the TGFBR2 E431K variant

To assess the impact of the TGFBR2 E431K variant on canonical TGF-β signalling, we performed *in vitro* assays in HEK293T cells transiently expressing wild-type (WT) *TGFBR2*, or the mutant construct. Quantitative RT-PCR analysis revealed that the E431K variant did not alter TGFBR2 mRNA levels compared with WT (Fig. S1). By contrast, immunoblotting revealed a marked decrease in TGFBR2 protein levels in E431K-expressing cells relative to WT, suggesting a decreased receptor stability (Fig. 4a). To assess whether the E431K variant affects the TGF-β signalling pathway, additional cell-based experiments evaluating SMAD2 activation and downstream transcriptional activation of target genes were performed. Following TGF-β1 stimulation (10 ng/mL, 1 h), SMAD2 phosphorylation was significantly reduced in E431K cells compared with WT (fold change: 2.867 ± 0.57 vs. 6.940 ± 0.094, *p < 0.01*) (Fig. 4b). In line with this, subcellular fractionation showed lower levels of nuclear phosphorylated SMAD2 accumulation in E431K-expressing cells relative to WT and pcDNA3.1+ controls, indicating decreased SMAD2 activation (Fig. 4c). Time-course analysis confirmed a persistent signalling defect at one, 6, and 9 hours post-stimulation (Fig. 4d). Finally, expression of TGF-β-responsive genes *SERPINE1* and *COL1A1* was significantly impaired in E431K-expressing cells (Fig. 4e). Collectively, these findings indicate that the E431K substitution promotes TGFBR2 degradation resulting in diminished canonical TGF-β transcriptional response demonstrated by reduced SMAD2 phosphorylation and impaired SMAD complexes nuclear translocation.

**Figure 4.**
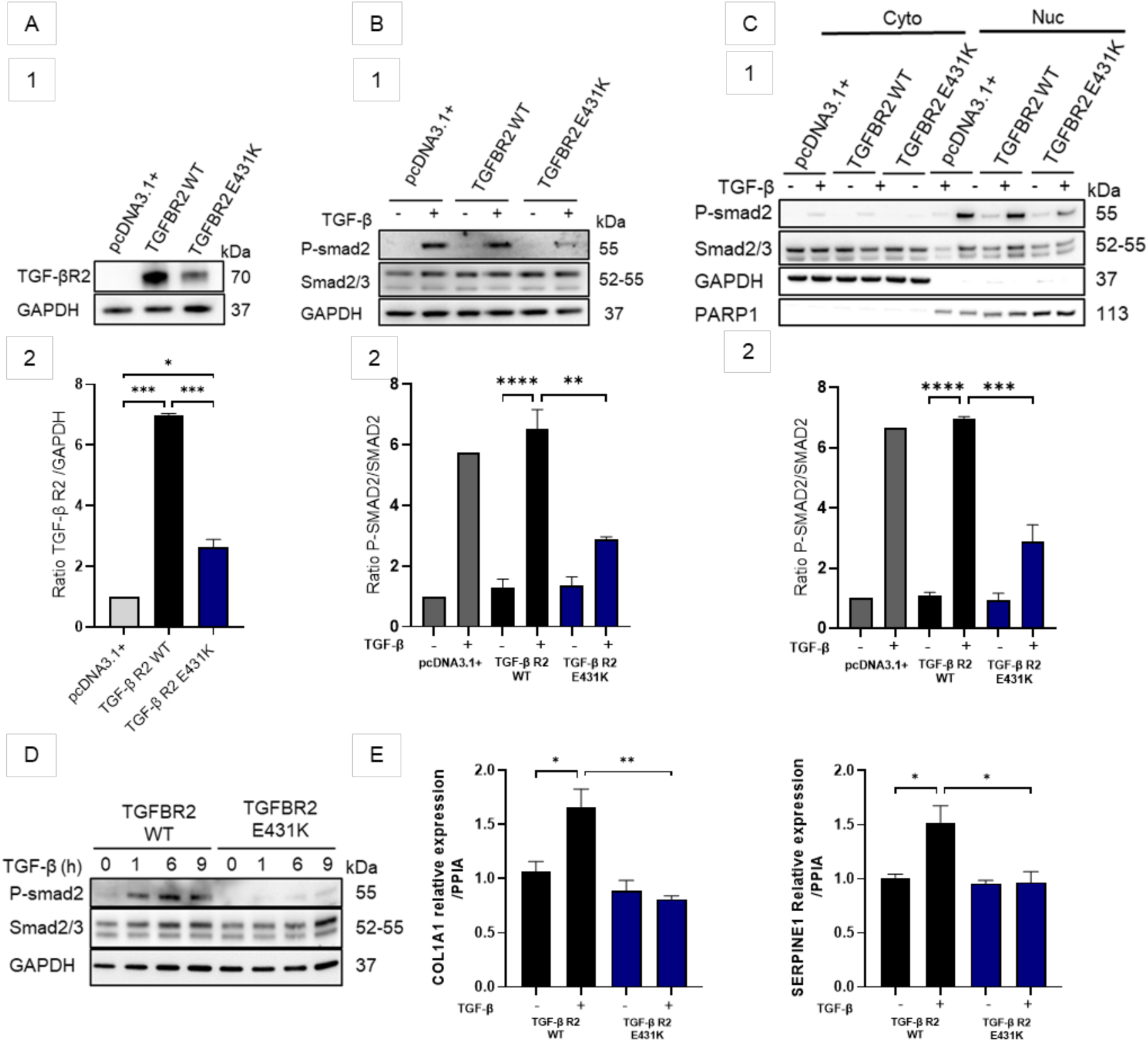
**TGFBR2 E431K disrupts the canonical TGF-β signalling pathway *in vitro.*** HEK293T cells were transiently transfected with 0.5 μg of pcDNA3.1+ empty vector, WT TGFBR2, or TGFBR2 E431K for 48 h and stimulated with TGF-β1 (10 ng/mL). Western blotting was used to assess (a) TGFBR2 abundance and SMAD pathway activation, including (b) whole-cell pSMAD2 and total SMAD2/3 and (c) nuclear pSMAD2 at 1 h post-stimulation, as well as (d) pSMAD2 levels at 1, 6, and 9 h post-stimulation. Downstream transcriptional responses (*SERPINE1*, *COL1A1*) were quantified by RT–qPCR. All experiments were performed in triplicate; Data are shown as mean ± Standard error of the mean (SEM), with statistical significance of *\*p < 0.05*, *\*\*p < 0.01*, *\*\*\*p < 0.001*, and *\*\*\*\*p < 0.0001*

## Discussion

Large-scale human genome sequencing initiatives have revealed extensive protein-altering genetic variations, of which only a minority are unambiguously deleterious usually by interfering with normal mRNA splicing or by introducing premature stop codons (Miosge et al. 2015). Missense mutations, which results in single amino acid changes in proteins, represents the most common class of protein-altering genetic variations and their functional impact on protein function largely depends on both the type of amino acid substitution and its protein context (Deák and Cook 2022; Sun et al. 2024). For example, the degree of evolutionary conservation of a given amino acid or its physico-chemical properties, including size, side chain polarity or flexibility, and its ability to form hydrogen bonds represent major features for optimal protein function (Ng and Henikoff 2003; Pace et al. 2014). Nevertheless, existing methods for interpreting the functional impact of missense variant suffer from inconsistent performance and poses a major challenge for their reliable use in clinical practice, with only an estimated 2% of missense variants that have been clinically categorized as pathogenic or benign (Fayer et al. 2021; Palafox et al. 2025). In this study, we identified in a patient with the clinical diagnosis of syndromic genetic aortopathy, two rare missense variants affecting *TGFBR2* and *FLNA* using whole-exome sequencing. The *TGFBR2* genetic variant was the most convincing candidate given the vascular phenotype of the patient overlapping with LDS and its absence from reference genomic databases, while the *FLNA* variant was ruled-out as disease causing given its presence in unaffected controls, patient gender and the absence of neurological manifestation typically associated with its loss-of-function. Indeed, mutations in the *FLNA* can cause X-linked dominant nodular ventricular heterotopia, a rare disease primarily affecting females and typically associated with a wide spectrum of connective tissue, skeletal, cardiovascular (including thoracic aortic aneurysm or dissection), and gastrointestinal disorders (Fergelot et al. 2012; Cannaerts et al. 2018).

To determine the functional impact of the TGFBR2 E431K variant, we used a combination of variant annotation, computational tools, and cell-based assays. E431 is located within the kinase domain, like the majority of missense variants reported to date in the most comprehensive compendiums of genetic variations relevant to human diseases such as ClinVar or HGMD. Importance of the E431 residue for TGFBR2 function is further suggested by its high conservation in vertebrates and the consistent presence of an acidic residue at a similar position in paralogous Ser/Thr kinases. Finally, many previously reported pathogenic or likely pathogenic variants in paralogs are also located in its close vicinity indicating that this residue is part of a functionally important protein site within the kinase domain (Loeys et al. 2005; Hara et al. 2019; Gupta et al. 2019). To better assess the functional consequence of the E431K variant, we took advantage of AlphaMissense, a recently developed cutting-edge AI model that integrates population frequency data along with structural and sequential contexts to predict the pathogenicity of every possible missense mutations in the human proteome (Cheng et al. 2023). In line with our findings, this algorithm confirms the functional importance of the kinase domain by showing high genetic constraint especially in the region surrounding the E431 residue and assigns to the E431K variant a high pathogenicity score. Then, to gain additional insights into the possible effects of this variant on *TGFBR2*, a high-quality homology-based model of this protein was generated using AlphaFold, a transformative AI-based tool with remarkable accuracy in *ab-initio* protein structure prediction (Abramson et al. 2024). Indeed, mapping genetic variations onto three-dimensional protein structures enables atomic-level analysis to assess the effects of amino acid substitution when protein is in its native and functional conformation. Interestingly, structural modelling positioned the E431 residue near the αF-helix, a core structural feature of the TGFBR2 kinase domain required to maintain its catalytic architecture and whose disruption has been shown to impair TGFBR2 function (Horbelt et al. 2010; Zhang et al. 2012). Moreover, the E431 residue is also adjacent to the Y424 residue, whose autophosphorylation is essential for TGFBR2 kinase activity (Lawler et al. 1997). Therefore, we hypothesize that substitution of the negatively charged glutamic acid with the positively charged lysine at position 431 in this critical protein region likely disturbs the electrostatic interactions and consequently the local conformation of TGFBR2 kinase domain. To support this hypothesis, we used the recently developed computational tool DDMut to predict change in Gibbs Free Energy upon E431K point mutation (Zhou et al. 2023). As expected, this algorithm forecasts that the glutamic acid to lysine substitution at position 431 of the TGFBR2 protein is sufficient to induce a significant destabilizing effect owing to the loss of polar and ionic interactions normally established with the Arg433 residue.

Previous reports have shown that disease-causing variants in *TGFBR2* result in abnormal canonical TGF-β signalling pathway *in vitro* (Horbelt et al. 2010; Cousin et al. 2017). Therefore, we assessed the effect of the E431K in human cells on SMAD signalling as a surrogate of canonical TGF-β pathway. Consistent with our *in silico* results, patient’s variant exhibited reduced SMAD2 activation and limited translocation into the nucleus as well as a reduced expression of SMAD target genes following TGF-β stimulation. This confirms the hypothesis that the glutamic acid to lysine substitution at position 431 induces TGFBR2 loss of function and consequently disrupts the downstream signalling cascade. Finally, to substantiate the predicted destabilizing effect induced by the E431K variant, we showed that mutant TGFBR2 protein expression is strongly reduced compared to wild type despite similar mRNA levels. Overall, these results highlight a charge-reversing amino acid substitution at a conserved position within the TGFBR2 kinase domain, which disrupts bonding networks, thereby reducing the stability of the receptor catalytic site and, consequently, impairing protein function.

In conclusion, we show the diagnostic utility of combining AI-driven protein modelling with *in vitro* cell-based assays to unambiguously established the molecular diagnosis of LDS and virtually all other rare mendelian genetic disorders in which aberrant TGF-β signalling plays a causative role. This study also further shows that protein destabilization by amino acid substitution play a prominent role in inherited human diseases.

## Materials and Methods

### Genetic Testing

Genomic DNA was extracted from EDTA-anticoagulated whole blood using the illustra Nucleon™ BACC3 Genomic DNA Extraction Kit (Cytiva; RPN8512), according to the manufacturer’s instructions. Exome sequencing was performed using the Illumina DNA Prep with enrichment (Illumina) and the actual sequencing was performed using the Novaseq 6000 Sequencing System (Illumina). Using this workflow, 97% of the targeted regions were covered at a depth of at least 20X. Data analysis, including base calling, de-multiplexing, alignment to the hg38 human reference genome (Genome Reference Consortium GRCh38), and variant calling, was performed using the Dragen v4.2.4 pipeline (Illumina). The TGFBR2 variant was confirmed by direct Sanger sequencing of the target region (Forward : ATGGGCCTCACTGTCTGTTT, Reverse : TGTCATTTCCCAGAGCACCA). Sequencing products were resolved by capillary electrophoresis on an ABI 3130XL automated fluorescent DNA sequencer, and sequences were analysed using SeqScape v2.5 (Applied Biosystems).

### Paralog Analysis

The paralog annotation approach was applied to assess the potential relevance of the E431K variant by comparing it with previously reported deleterious substitutions in TGFBR2 and its paralogous proteins. Multiple sequence alignment (MSA) of paralogs for the gene ENSG00000163513 (transcript: ENST00000295754; GRCh37.p13) was obtained from Ensembl (Cunningham et al., 2015). The alignment was generated using Clustal Omega (Sievers et al., 2011; McWilliam et al., 2013) with default parameters. Reported pathogenic variants in paralogous genes were extracted from ClinVar (Landrum et al., 2014, 2018) and the Human Gene Mutation Database (HGMD Professional, latest release; Stenson et al., 2020).

### Structure prediction using AlphaFold3

Human TGFBR2 (Uniprot P37173, https://www.uniprot.org/uniprotkb/P37173/entry) and TGFBR2 E431K variant sequences were submitted to AlphaFold3 server (https://alphafoldserver.com/) with standard parameters ^1^. Quality of the predicted structures from AlphaFold3 was asses using the pLDDT and pTM confidence scores. A pTM score above 0.5 indicated that the overall predicted fold for the complex is likely similar to the true structure. pLDDT is a per-atom confidence estimate on a 0-100 scale where a higher value indicates higher confidence. (Fig. S5).

AlphaMissence predictions data for the TGFBR2 were extracted from the AlphaFold Protein Structure Database (https://alphafold.com/). AlphaMissense is an AI model that builds on Google DeepMind’s AlphaFold2 to classify ‘missense’ mutations in proteins as either ‘likely pathogenic’, ‘likely benign’ or ‘uncertain’ with a score from 0 to 1^2^. Predicted structures were visualized using ChimeraX (version 1.10.1) ^3^.

### Cell Culture and transient transfection

Human embryonic kidney HEK293T cells (ATCC® CRL-3216™) were maintained in Dulbecco’s Modified Eagle Medium (DMEM; Gibco™) supplemented with 10% fetal bovine serum (FBS; Gibco™) and 1% penicillin–streptomycin (Gibco™). Cells were cultured at 37 °C in a humidified incubator with 5% CO₂. Subculturing was performed at 70-80% confluence using 0.05% trypsin–EDTA (Gibco™).

HEK293T cells were seeded at 3×10^^5^ cell per well and transiently transfected for 48 h with pcDNA™3.1(+) plasmids (Thermo Fisher scientific, V79020) encoding wild-type TGFBR2 (TGFBR2 WT) or the c.1291G>A variant (TGFBR2 c.1291G>A, p.Glu431Lys, E431K), or with the empty pcDNA3.1(+) vector as control using Lipofectamine™ LTX Reagent with PLUS™ Reagent (Thermo Fisher scientific, 15338100). Insert sequence and orientation were validated by Sanger sequencing. Following transfection, cells were serum-starved for 16 h in serum-free medium and subsequently stimulated with recombinant human TGF-β1 (10ng/mL) for 1, 6, or 9 h prior to downstream analyses.

### Western Blotting

Treated cells were collected and lysed in radioimmunoprecipitation assay (RIPA) buffer supplemented with a protease and phosphatase inhibitor cocktail (Roche), or subjected to nuclear and cytoplasmic fractionation using the NE-PER™ Nuclear and Cytoplasmic Extraction Reagents kit (Thermo Fisher Scientific). Protein concentrations were determined using the Thermo Scientific™ Pierce™ BCA Protein Assay Kits (Thermo Fisher Scientific), and equal amounts of protein (10 µg per sample) were denatured, resolved on 4–12% Bis-Tris polyacrylamide gels (NuPAGE™, Thermo Fisher Scientific), and transferred onto nitrocellulose membranes. Membranes were blocked with 5% non-fat dry milk in Tris-buffered saline (TBS) for 1 h at room temperature, followed by overnight incubation at 4 °C with the following primary antibodies: TGF-β Receptor II (E5M6F) (1:1,000; Cell Signaling #41896), total SMAD2/3 (1:1,000; Cell Signaling, #8685), phospho-SMAD2 (1:1,000; Cell Signaling, #3108), PARP1 (E102) (1:5,000; Abcam, #32138), and GAPDH (1:1,000; Santa Cruz Biotechnology, sc-32233). The next day, membranes were incubated with horseradish peroxidase (HRP)-conjugated secondary antibodies for 1 h at room temperature. Protein bands were visualized using Amersham ECL Select™ reagent (Cytiva) and imaged with a GE Image Quant LAS-4000 system. Band intensities were quantified using ImageJ software.

### RNA Extraction and Quantitative Real-Time PCR (qPCR)

Total RNA was extracted from cultured cells using the RNeasy Mini Kit (Qiagen™). RNA concentration and purity were determined by spectrophotometry (NanoDrop™, Thermo Fisher Scientific™). For reverse transcription, 1 µg of total RNA was converted into cDNA using the High-Capacity cDNA Reverse Transcription Kit (Applied Biosystems™). Quantitative PCR was performed using TaqMan™ Gene Expression Assays (Applied Biosystems™) on a QuantStudio™ real-time PCR system (Applied Biosystems™). Each reaction was carried out in triplicate in a 20 µL volume containing TaqMan™ Universal PCR Master Mix, cDNA template, and gene-specific TaqMan™ probes (TGFBR2 Hs00234253_m1 COL1A1 Hs00164004_m1, SERPINE-1 Hs01126607_g1. PPIA (Hs99999904_m1) was used as an endogenous control. Relative gene expression levels were calculated using the 2^^−ΔΔCt^ method.

### Statistical analysis

Statistical analyses were performed using GraphPad Prism software. Results are given as mean ± SEM. T-test was used for single comparisons; one-way ANOVA followed by Tukey post hoc test was used for multiple comparisons. p-value less than 0.05 was considered statistically significant.

## Data Availability

Data produced in the present study are available upon reasonable request

## CRediT authorship contribution statement

Nihad Boukrout: Writing – original draft, Methodology, Formal analysis, Data curation. Clément Delage: Formal analysis, Data curation. Thomas Comptdaer: Methodology, Formal analysis, Investigation. Wendy Arondal: Formal analysis. Amine Jemel: Resources. Nadia Azabou: Resources. Mouna Bousnina: Resources. Molka Mallouki: Data curation. Naoual Sabaouni: Investigation. Rihab Arbi: Resources. Salma Kchaou: Resources. Haithem Ammar: Resources. Saoussen Hantous-Zannad: Resources. Houweyda Jilani: Resources, Formal analysis. Yasmina Elaribi: Resources. Lamia Benjemaa: Resources. Cynthia Van der Hauwaert: Formal analysis. Romain Larrue: Writing – review & editing. Meyling Cheok: Formal analysis. Michaël Perrais: Formal analysis, Writing – review & editing. Bruno Lefebvre: Methodology, Formal analysis, Writing – review & editing. Christelle Cauffiez: Project administration, Writing – review & editing. Nicolas Pottier: Writing – review & editing, Supervision, Conceptualization.

## Ethics declaration

This study complies with French laws and has been approved by the Institutional Review Board of Centre Hospitalier Universitaire de Lille (France). All participants provided written informed consent for genetic analysis and publication of this study in line with institutional guidelines and the Declaration of Helsinki and Istanbul. The DNA collection is officially registered with the “Ministère de l’Enseignement Supérieur et de la Recherche (Paris, France)” under the number: DC-2008–642.

## Data availability

The data (clinical history and pedigree) are available upon request from the authors.

## Conflict of interests

The authors declared no competing interests.

## Acknowledgements

We thank the patient for his participation in this study.

## Notes

### Competing Interest Statement

The authors have declared no competing interest.

### Funding Statement

This study did not receive any funding

### Author Declarations

This study complies with French laws and has been approved by the Institutional Review Board of Centre Hospitalier Universitaire de Lille (France). All participants provided written informed consent for genetic analysis and publication of this study in line with institutional guidelines and the Declaration of Helsinki and Istanbul. The DNA collection is officially registered with the Ministere de lEnseignement Superieur et de la Recherche (Paris, France) under the number: DC2008642.

